# Dietary plant diversity predicts early life microbiome maturation

**DOI:** 10.1101/2025.02.28.25323117

**Authors:** Teresa K. McDonald, Ammara Aqeel, Ben Neubert, Anna Bauer, Sharon Jiang, Olivia Osborne, Nolan Ives, Danting Jiang, Filemon Bucardo, Lester Gutiérrez, Luis Zambrana, Kirsten Jenkins, Jennifer Gilner, Javier Rodriguez, Amanda Lai, Jonathan P. Smith, Rinn Song, Kazi Ahsan, Sheraz Ahmed, Sanam Iram Soomro, Fayaz Umrani, Michael Barratt, Jeffrey I. Gordon, Asad Ali, Najeeha Iqbal, Jillian H. Hurst, Victoria Martin, Wayne Shreffler, Qian Yuan, Joe M. Brown, Neeraj K. Surana, Samuel Vilchez, Sylvia Becker-Dreps, Lawrence A David

**Affiliations:** Department of Molecular Genetics and Microbiology, Duke University School of Medicine, Durham, North Carolina, USA; Harvard Medical School, Boston, Massachusetts, USA; Food Allergy Center, Massachusetts General Hospital, Boston, Massachusetts, USA; Division of Pediatric Gastroenterology and Nutrition, Massachusetts General Hospital for Children, Boston, Massachusetts, USA; Division of Pediatric Allergy and Immunology, Massachusetts General Hospital for Children, Boston, Massachusetts, USA; Department of Family Medicine, University of North Carolina at Chapel Hill, Chapel Hill, North Carolina, USA; Department of Epidemiology, University of North Carolina at Chapel Hill, Chapel Hill, North Carolina, USA; Department of Microbiology and Immunology, University of North Carolina Chapel Hill, Chapel Hill, North Carolina, USA; Universidad Nacional Autónoma de Nicaragua León, León, Nicaragua; Department of Pediatrics, Division of Infectious Diseases, Duke University School of Medicine, Durham, North Carolina, USA; Department of Obstetrics and Gynecology, Division of Maternal-Fetal Medicine, Duke University School of Medicine, Durham, North Carolina, USA; Department of Pediatrics, Children’s Clinical Research Unit, Duke University School of Medicine, Durham, North Carolina, USA; Department of Environmental Sciences and Engineering, Gillings School of Global Public Health, University of North Carolina at Chapel Hill, Chapel Hill, North Carolina, USA; The Aquaya Institute, San Anselmo, California, USA; Duke Microbiome Center, Duke University School of Medicine, Durham, North Carolina, USA; Department of Health Policy and Management, Yale School of Public Health, New Haven, Connecticut, USA; Division of Global HIV and Tuberculosis, U.S. Centers for Disease Control and Prevention, Atlanta, Georgia, USA; Oxford Vaccine Group, Department of Pediatrics, University of Oxford, Oxford, UK; The Edison Family Center for Genome Sciences and Systems Biology, Washington University School of Medicine, St. Louis, MO 63110 USA; Newman Center for Gut Microbiome and Nutrition Research, Washington University School of Medicine, St. Louis, MO 63110 USA; Aga Khan University, Medical College, Pakistan; Department of Pediatrics, Duke University School of Medicine, Durham, North Carolina, USA; Centro de Investigación de Enfermedades Tropicales (CIET). Facultad de Microbiología. Universidad de Costa Rica, San José, Costa Rica

## Abstract

Despite established links between the infant gut microbiome and health, how complementary feeding shapes colonization remains unclear. Using FoodSeq, a DNA-based dietary assessment technique, we surveyed food intake across 729 children (0-3 y) from North and Central America, Africa, and Asia. We detected 199 unique plant food sequences, with only eight staples shared across all countries. Despite this global variation, early-life diets followed a common trajectory: weaning stage emerged as the dominant dietary signature across populations. Crucially, dietary diversity did not correlate with gut microbial diversity. Instead, dietary diversity and weaning stage specifically predicted the abundance of adult-associated bacteria. Our findings support a two-stage maturation model: an initial milk-dominated phase, followed by a diet-responsive phase that yields adult-like communities. Monitoring and promoting plant dietary diversity may thus support timely microbiome maturation worldwide.

## Introduction

In the first years of life, the human gut transitions from a sterile state to a diverse microbial ecosystem. During this transition, the gut microbiome transforms from an ‘immature’ state -- characterized by a limited set of milk-adapted taxa -- to a ‘mature’ adult-like community harboring fiber-degrading taxa and a diversity of other commensal species^1–3^. Proper microbial succession is fundamental to healthy metabolism, immune development, and disease resistance. Indeed, disruptions in early life colonization and microbiome maturation increase risk of conditions such as obesity^4^, allergy^5^, and Type I diabetes^6^. Consequently, the first years of life represent a critical window when microbiome development can have lifelong health effects.

Despite the importance of the gut microbiome in healthy infant development, the specific factors guiding early microbiome colonization remain less clear. Well-established influences include antibiotic exposure, which reduces bacterial diversity and stability; caesarean delivery, which alters the initial microbial repertoire; and formula feeding, which lowers *Bifidobacterium* abundance yet increases overall microbial diversity^7^. Another pivotal factor is cessation of breastfeeding^3,2,8^, which triggers a decline in the abundance of milk-adapted microbial taxa.

One area of microbiome development, that remains underdeveloped, however, is the precise role that complementary feeding plays in microbial succession^1^. While diet is known to reproducibly alter the gut microbiome in adults^9^, few studies have found evidence directly linking specific dietary factors to microbiome maturation by concurrently measuring diet and the microbiome in young children. Exceptions have either tested narrow food interventions^10–13^ or were conducted within a single cultural and dietary context^14–17^. Other studies have measured microbiome composition across more diverse dietary traditions^8,18^; still such studies have been limited in sample size or have often lacked detailed dietary data, leading comparisons to be on population-level dietary patterns rather than individual-level diet metrics.

A major barrier to understanding the relationship between diet and microbiome maturation is the challenge of accurately characterizing infant diet. Surveys, the gold standard and traditional approach for collecting dietary data, are labor-intensive^56^ require baseline levels of literacy^51^, incompletely capture minority or ethnic foods^52–54^, and are vulnerable to social desirability bias^19,20^. Dietary assessment in children poses additional obstacles, as it relies on secondhand reporting by parents and guardians who may not observe all meals when children are under the supervision of other caregivers. In the United States, for instance, a quarter of 2-5 year olds’ energy intake is consumed outside the home^21^. Such incomplete observation contributes to reporting bias exemplified by validation studies with doubly-labeled water that show infant dietary intake is overestimated by 7-29%^22–27^. Finally, the technical challenges with infant dietary assessment only multiply in multi-country settings, as assessments must be adapted for each different geographic, cultural, and linguistic setting^28^ For example, there are over 100 versions of a standardized survey for comparing population-level global diets^29^. In concert, these barriers have prevented systematic investigation of the relationship between early life microbiome development and infant diet across diverse human populations.

### Remarkable diversity in early-life diets worldwide

To perform a global survey of infant microbiome development and diet, we assembled a meta-cohort from existing longitudinal studies and population health surveillance surveys of 1036 fecal samples from 729 children aged 0-3 years from five countries across multiple continents: the USA (n=144), Nicaragua (n=457), Cambodia (n=178), Kenya (n=228), and Pakistan (n=29). These sites were selected to encompass wide variation in geography, socioeconomic status, and dietary traditions, as well as varying burdens of infectious diseases common in early childhood. Stool samples and dietary information were collected; further details on cohort characteristics and methodologies are provided in the methods and Table S1.

We applied FoodSeq, an objective DNA sequencing-based dietary assessment^30^, to our cohort. FoodSeq is an amplicon sequencing-based technique that sequences the leucine gene (*trnL*) from plant plastids or the 12S rRNA gene from animal mitochondria. By mapping *trnL* and 12S sequencing reads to a curated reference database, FoodSeq identifies and quantifies foods of plant and animal origins at a taxonomic level. Unlike traditional dietary surveys, FoodSeq provides objective measurements that are independent of caregiver reporting and remain consistent across varying income levels, health conditions, and infant feeding practices. The diet data generated by FoodSeq also has the advantage of being temporally synchronized with corresponding microbiome data, as both sets of genomic analyses can be generated from the same fecal sample.

FoodSeq revealed extensive variety and breadth in early life diets across our global cohort (Fig. 1). With plant FoodSeq, we detected 113 species and 86 assigned sequence variants (ASVs) that matched at a higher level (e.g. genus), encompassing 389 potential dietary plant species. Of the plant ASVs, 42% were detected exclusively in a single country (Fig. S1A), with unique country-specific foods ranging from longan and lotus root in Cambodia, to jute mallow and cassava in Kenya, to raspberry in the USA (Fig. 1A). By contrast, just eight global staples^31^ showed consistent prevalence in all five countries (detected in >5% of each country’s samples): wheat, corn, rice, banana/plantain, tomatoes and other nightshades (potato, etc.), mangifera (mangos), and alliums (onion, leek, garlic, chives) (Fig. 1A, foods in bold).

**Fig. 1:**
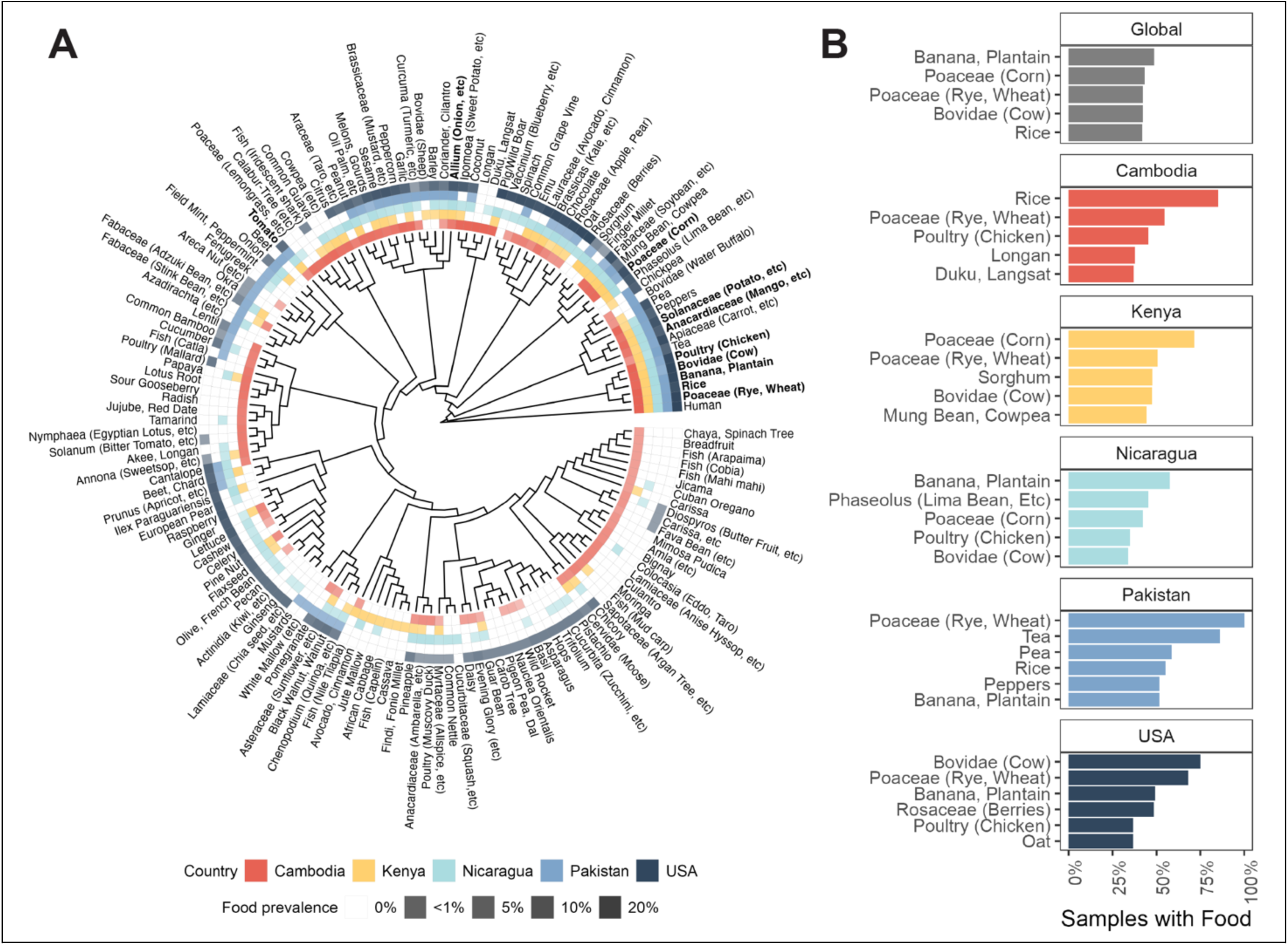
Early life diet is diverse and varies globally. (A) Dendrogram of food prevalences. Rare foods (<1% frequency in at least one country) were filtered out prior to plotting. Sequences that matched to a higher taxonomic level than species are labeled with the lowest taxonomic level assigned (plants) or a representative taxonomic level (animals), and then a common name of a food that sequence matches. Sequences with multiple foods or food names associated with them are labeled with ‘etc’. The 8 plant and 2 animal foods detected in at least 5% of samples in all 5 countries are labeled in bold. (B) Top 5 most prevalent foods in each country.

Despite the breadth of plant species in our global survey, principal component analysis (PCA) revealed that the primary axis of dietary variation, PC1, was likely driven by the overall presence of plant matter in the diet rather than specific dietary composition (Fig. 2A-B). Unlike subsequent PCs, which displayed mixed positive and negative loadings indicative of tradeoffs in plant species abundance, PC1 exhibited exclusively positive loadings for common foods (Fig. 2C). This pattern suggests that PC1 captures the extent of plant consumption rather than regional dietary differences. Supporting this interpretation, PC1 was strongly correlated with both infant age (Fig. 2D, Spearman’s *ρ* =0.25, p<0.001) and overall plant FoodSeq richness (pFR, the number of unique sequences assigned to our human foods database in each sample; Fig. 2E, *ρ* =0.69, p<0.001). These associations are consistent with the expected trajectory of weaning as infants gradually incorporate more variety of solid foods into their diets as they mature. Direct comparisons of dietary diversity and age confirmed our interpretation, showing that pFR consistently increased across the first year of life until stabilizing (Fig. 2G; Spearman’s *ρ* =0.27, p<0.001). Taken together, these findings argue that FoodSeq effectively captures the dietary shifts associated with weaning.

**Fig. 2:**
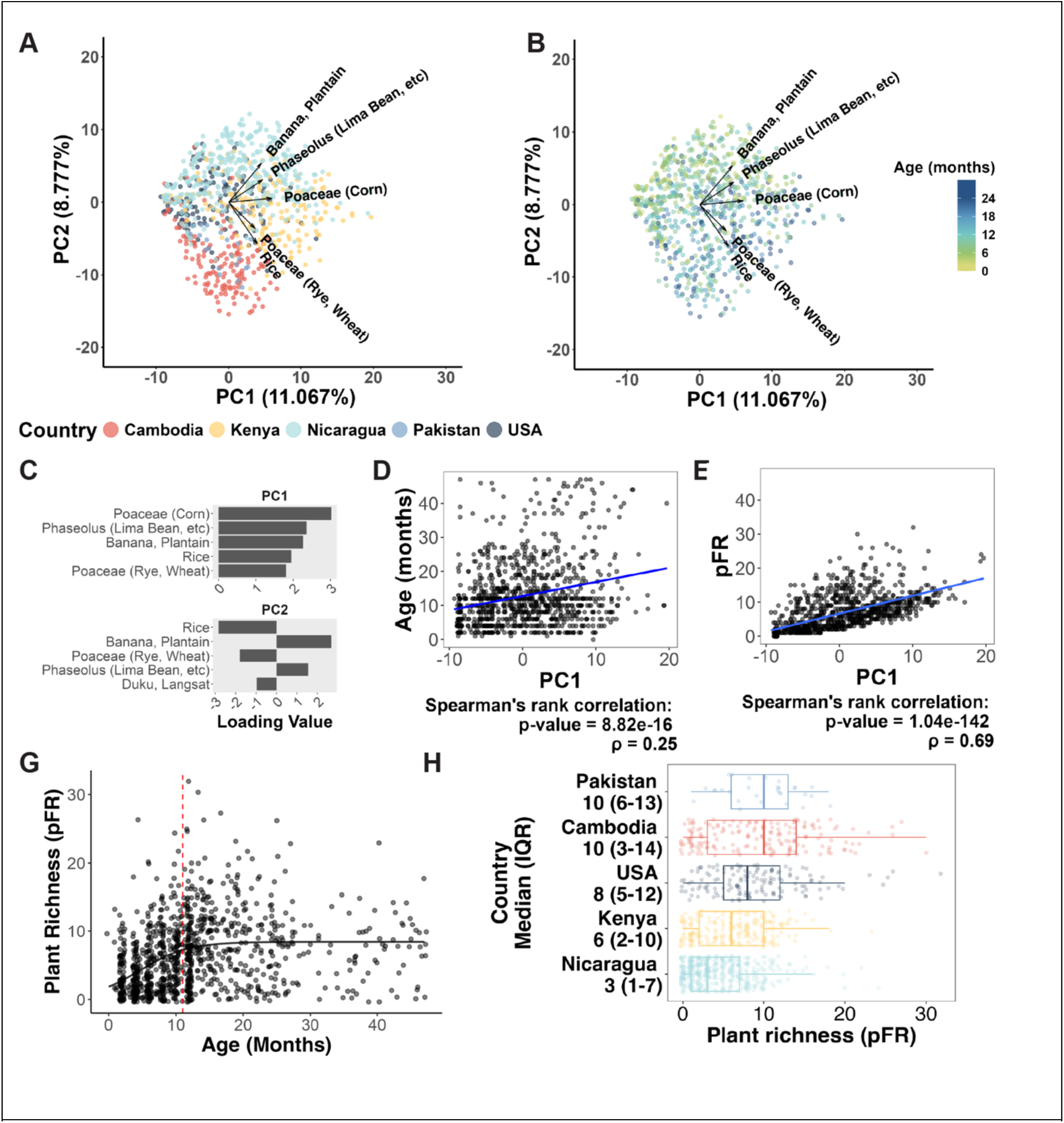
Dietary variation across age and country. **(A)** Plant FoodSeq PCA colored by country. Country accounted for 15% of variation, age accounted for 0.6% variation, and dietary diversity accounted for 5% of variation. (PERMANOVA, p-value <0.001 for all). **(B)** PCA colored by age **(C)** PCA loadings. **(D)** Correlation between age and PC1. (E) Correlation between pFR and PC1. **(G)** pFR over time. A logistic line (black) was fit to the data. The red dashed line represents the plateau point at which the increase in pFR decelerates, calculated as the x value at which the first derivative of the line reaches 50% of its maximum. **(H)** pFR by country.

While weaning stage dominated the first PC, the second PC revealed country-specific dietary signatures partitioned by their relative intakes of regional staple plants like rice (enriched in Cambodia), banana/plantain (enriched in Nicaragua), or sorghum and millet (enriched in Kenya). Cross-country differences also emerged in dietary diversity more broadly, with median pFR ranging from 4 in Nicaragua to 10 in Cambodia (Fig. 2H). Such values aligned with previously reported food variety scores, which found mean food variety ranges from 5.5 to 10 among children from 6-24 months in Africa and Central America^32–34^. The timing and rate of dietary diversification also varied by country. For instance, the diets of Cambodian children diversified early and rapidly until pFR plateaued at 8 months of age, whereas US infants showed a more gradual rise in pFR until 13 months (Fig. 3B).

**Fig. 3.**
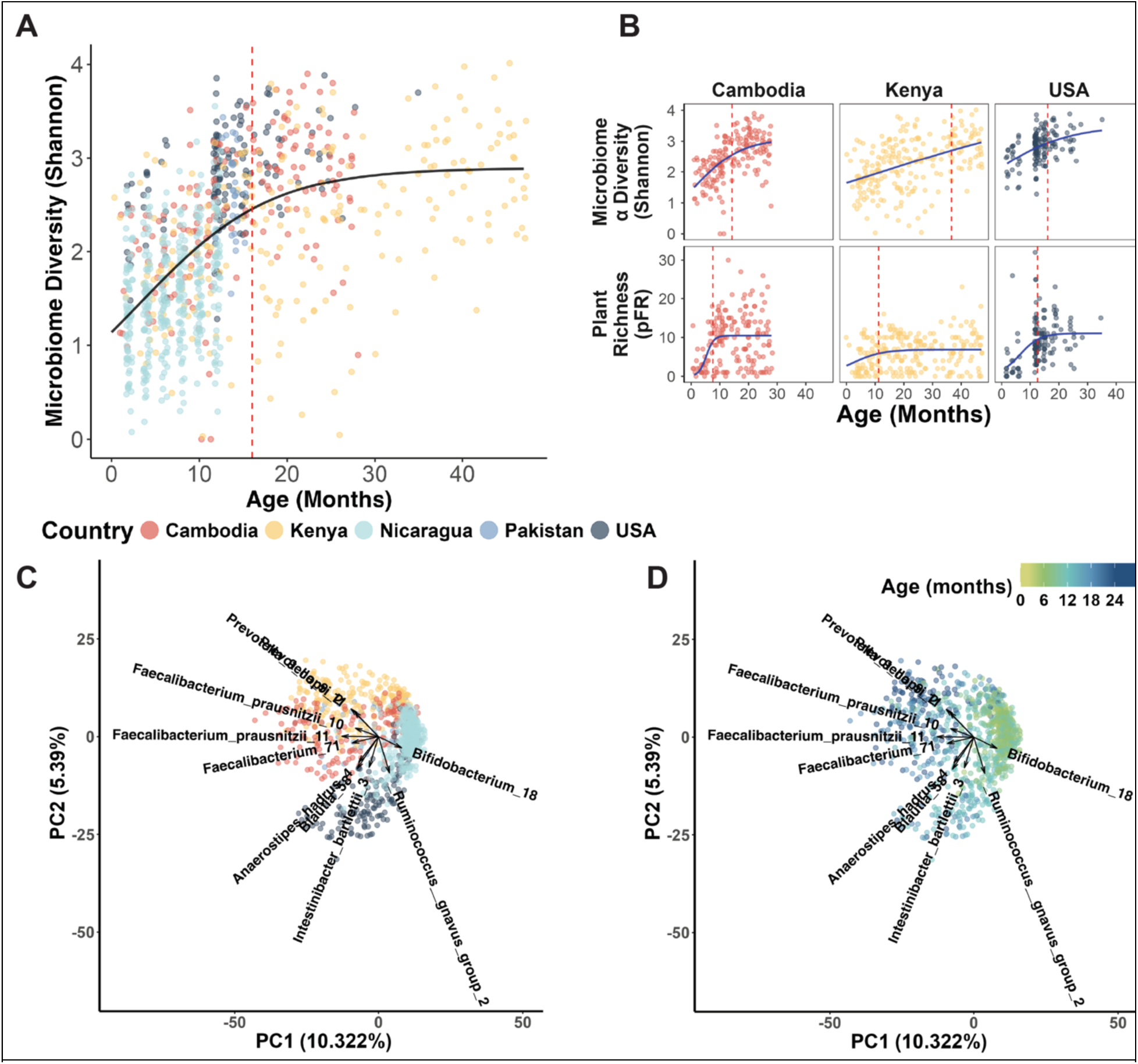
The early life microbiome varies primarily by geography and age. **(A)** Microbiome diversity increases with age. A logistic line (black) was fit to the data. The red dashed line represents the point at which diversity plateaus with respect to age, defined as the point at which the first derivative of the line reached 50% of its maximum. **(B)** Dietary diversity plateaus prior to microbiome diversity. A logistic line (black) was fit to the data and the plateau point (red dashed line) calculated as in (A). **(C)** PCA of 16S rRNA sequencing data colored by country. **(D)** PCA of 16S data colored by age in months.

We note that in contrast to the diversity of plant-associated species and dietary patterns observed in our cohort, we detected a more limited breadth of dietary animals. A total of 28 dietary animal species were detected. Observed animals included widely utilized livestock such as pig, chicken, and cow, as well as more region-specific animals including water buffalo, the primary source of milk in Pakistan, and various fish species in Cambodia. Overall, however, 41% of infant samples had no non-human animal DNA detected, and only 33% had more than one animal species detected. Only the US and Cambodia had a non-zero median count of animal species present (Fig. S1B). The limited detection of dietary animals was consistent with the higher cost of animal-based foods and the majority of human animal-based foods originating from cattle, poultry, and pigs^28^ for the locations of infants sampled here. Given the limited detected range of animal species, the distinct nutritional characteristics of different animal products from the same species (beef, dairy), as well as the known importance of nutrients like fiber for the gut microbiome, we focused our subsequent analyses on plant-based foods and gut microbiome development.

### Patterns of microbiome maturation

Prior to analyzing the relationship between early-life dietary and microbiome development, we first confirmed that our cohort followed well-established patterns of early-life gut microbiome maturation^3^. Microbiome alpha-diversity increased steadily from birth (Fig. 3A) through age two, regardless of country (Fig. 3B). In terms of inter-individual variation (beta-diversity), both country of origin (Fig. 3C) and age (Fig. 3D) were significant factors (country R^2^=0.4675, p=0.001, age R^2^=0.4721, p=0.001; EnvFit on Unweighted Unifrac. Fig. S2A). In cohorts where data were available on breastfeeding status (USA, Nicaragua, Cambodia) and birth mode (USA, Nicaragua), both factors were associated with microbiome composition. (Breastfeeding: R^2^= 0.1377, p=0.001; Birth mode: R^2^= 0.0088, p=0.014, Envfit on unweighted Unifrac, controlling for age and country). Thus, consistent with previous literature, we observed both increased diversification of the microbiome with age, as well as notable roles for breastfeeding status and birth mode in shaping microbiome structure in our cohort^35^.

The specific bacterial taxa observed in our infant cohort also aligned with prior studies^1–3,8^, as hierarchical clustering revealed a microbial succession pattern (Fig. S3A). We identified an early-life cluster (Cluster 9), enriched in *Bifidobacterium* and *Streptococcus,* and a ‘transitional’ cluster (Cluster 1), abundant at ages 12-18 months, characterized by plant degraders, *Blautia* and *Ruminococcus,* as well as the probiotic genus *Ligilactobacillus* (Fig. 4A).

**Fig. 4.**
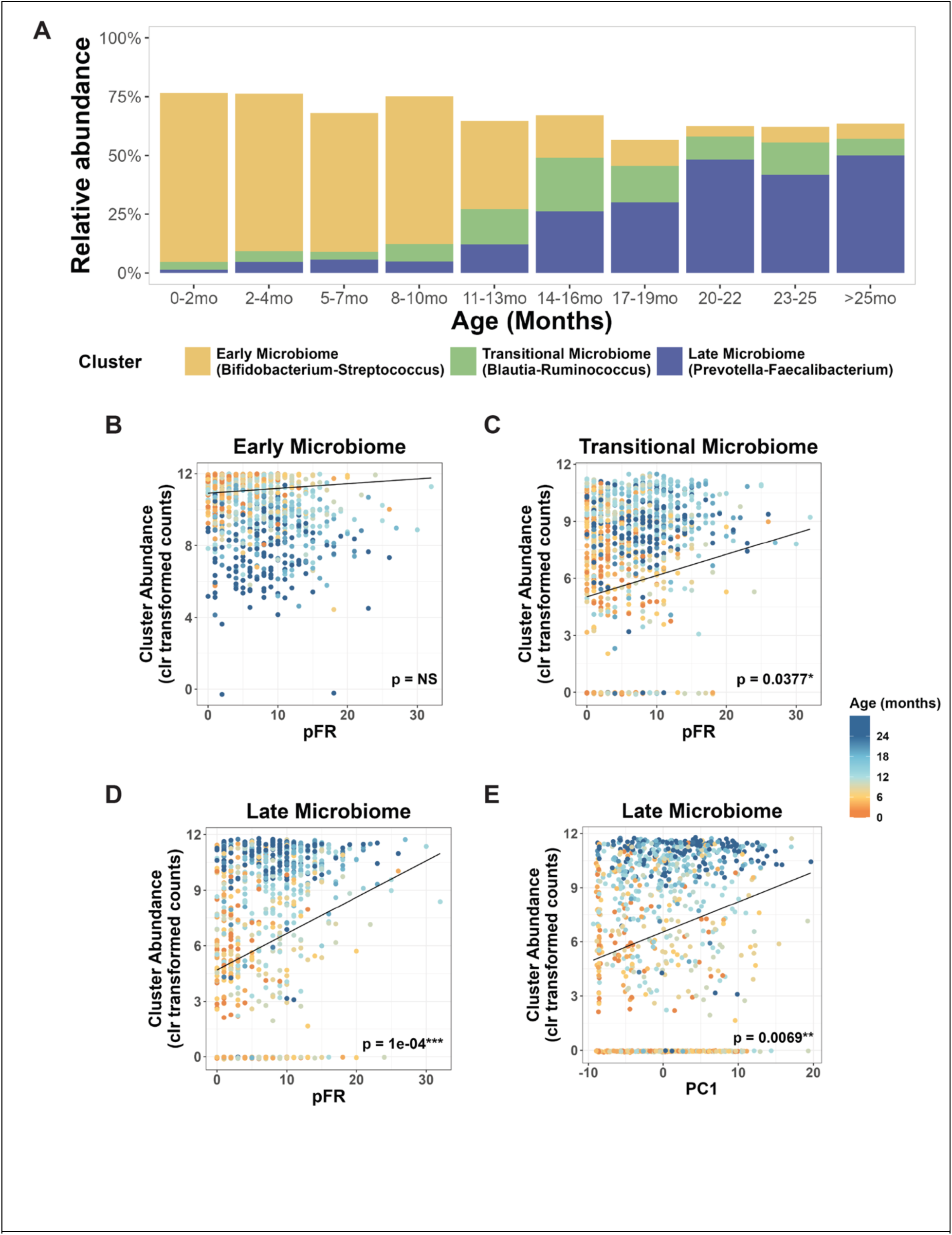
Plant dietary diversity is correlated with the abundance of mature-microbiome marker bacteria. **(A)** Cluster abundance over time. **(B)** The abundance of the ‘Early Microbiome’ cluster, plotted against plant richness, **(C)** The abundance of the ‘Transitional Microbiome’ cluster, plotted against plant richness. (**D-E)** The abundance of the ‘Late Microbiome’ cluster, plotted against plant richness **(D)** and PC1 **(E).** For B-E, lines plot the coefficient derived from a generalized linear model (GLM) controlling for age and country.

Following this transition, a late-microbiome cluster (Cluster 5) emerged between 21-36 months that more closely resembled the adult microbiome, featuring *Bacteroides vulgatus* and *Faecalibacterium prausnitzii,* which ferment plant fiber into butyrate and other short chain fatty acids, and *Prevotella copri,* a core fiber-metabolizing taxon^36^ (Fig. 4A). In parallel, we constructed a Random Forest model that successfully predicted infant age given input microbiome data. The RF model identified *Faecalibacterium* and *Bifidobacterium* as top predictor variables, mirroring both our hierarchical clustering results and previously published models of infant microbiome development^1^ (Fig.S4).

### Dietary diversity does not predict microbiome diversity

Having found that infant microbiota in our cohort matured as expected from previous studies, we next compared patterns of dietary maturation to gut microbiome maturation.

Surprisingly, we did not see consistent evidence that dietary diversity predicted gut microbial diversity. Microbial alpha diversity continued to increase until at least 14-16 months of age (Fig. 3A) well after dietary diversity (as measured by pFR) had plateaued (Fig. 3B; 2G). We did observe associations between dietary diversity and microbiome diversity within several countries but associations were not consistent; for example, pFR correlated with microbiome Shannon diversity in Kenya and the United States, whereas in Nicaragua, the relationship was reversed (Fig. S2C). Further, when we constructed global models controlling for age and country, pFR did not correlate wtih overall microbiome diversity measures (Shannon diversity and 16S richness both non-significant (Fig. S2D, F). Thus, although children began eating complex diets soon after introduction of solid foods, the gut microbiome continued to diversify even after dietary complexity had been reached.

### Diet is associated with later stage microbes during gut colonization

We next hypothesized that although dietary diversity may not directly correlate with overall microbial diversification, plant richness and weaning stage would be associated with microbiome maturity. Indeed, despite substantial geographic variation in diet, we found pFR was positively correlated with the ‘Transitional Microbiome’ cluster, comprised of *Blautia* and *Ruminococcus*, and the ‘Late Microbiome’ cluster, comprised of *Bacteroides*, *Faecalibacterium*, and *Prevotella*, even after controlling for age and country (Fig. 4C, D). Likewise, PC1 was positively correlated with ’Late Microbiome’ cluster abundance (Fig. 4E).

Given that we sourced fecal samples across multiple study sites, we re-fitted all models while (i) adjusting for diarrhea, weight-for-height z-score, RUTF intake, and sequencing depth, and (ii) adding participation in the Cambodian nutrition-education intervention. None of these adjustments altered the direction or significance of the associations between plant richness (pFR) and the Transitional or Late microbiome clusters, nor did they create any association with Shannon diversity (Fig. S5,6).

Ultimately, as increased dietary diversity (i.e., pFR) and weaning stage (i.e., PC1) both reflect complementary feeding and the introduction of an adult-like diet, our findings support the hypothesis that introduction of an adult-like diet stimulates maturation toward an adult-like microbiome. Notably, the ’Early Microbiome’ cluster did not correlate with dietary diversity (Fig. 4B) or PC1, consistent with the hypothesis that milk consumption, rather than solid food introduction, primarily shapes gut microbial ecology prior to weaning. These results suggest that the transition to an adult-like microbiome specifically is driven by the cumulative introduction of diverse solid foods, but that other transitions within the microbiome maturation process, such as overall microbiome diversification, are not driven by solid food diversity. These transitions are furthermore unlikely to be driven by consumption of particular foods or specific dietary patterns, as we observed conserved microbiome microbial responses to diversity across countries with highly unique and non-overlapping diets. More broadly, our findings point to a developmental sequence wherein milk consumption governs early microbiome composition, while the subsequent introduction of solid plant foodsdrives the transition toward a mature gut microbial community.

## Discussion

In concert, our findings here refine existing models linking weaning to microbiome development in the infant gut. Our data do not reveal simple linear relationships between dietary and microbiome diversity across early life. This negative finding, however, is consistent with our increasing understanding that microbial ecosystem diversity does not necessarily predict community function^37,38^. Instead, our data support a two-stage developmental model: an early phase governed by milk consumption, followed by a maturation phase shaped by dietary diversity. In this second stage, plant dietary diversity and the child’s physiologic age together predict the colonization by select groups of gut bacterial taxa associated with adult-like microbiome function. Notably, despite the broad range of complementary feeding patterns across our geographically diverse cohorts, we observed similar successional trends, underscoring the functional redundancy^39^ that gut microbes can share, regardless of environmental differences.

Early life nutrition remains one of the most critical challenges in global health, with lasting consequences for child development and lifelong health outcomes^40^. Our findings confirm that adequate and diverse plant consumption during the complementary feeding period foster maturation of the gut microbiome toward an adult-like state rich in fiber-degrading bacteria, extending the known benefits of dietary diversity beyond micronutrient adequacy^41,42^ and food allergy prevention^43^. Importantly, we show that simply counting the number of distinct plant foods effectively measures this microbial transition, potentially offering a more accessible and practical complement to widely used dietary indices such as the WHO minimum dietary diversity score (MDD), which assesses diversity as consumption across at least five of eight food groups.^44^ Implementing straightforward, ‘pro-gut’ complementary feeding guidance based on overall plant dietary richness could enable easier and more precise monitoring by caregivers and health workers across diverse settings, ultimately supporting appropriate microbiome maturation among infants worldwide.

## Materials and Methods

### Study populations and stool sampling

#### Cambodia

Cambodian samples were obtained as extracted DNA from stool samples collected as part of the Integrated Nutrition, Hygiene and Sanitation Project (NOURISH), conducted across three rural provinces^45^. The NOURISH study involved both nutrition and sanitation interventions. The nutrition intervention specifically consisted of caregiver nutrition education, direct cash payments contingent upon regular health visit attendance, and provision of two $5 food vouchers. Of the Cambodian samples in our study, 101 (56.7%) originated from children enrolled in either of the nutrition intervention arms (arm 1: nutrition intervention only; arm 2: nutrition intervention+sanitation). Our selection prioritized samples from children who had not experienced diarrhea or fever within the week preceding sample collection; however, 14 samples came from children who had recently experienced diarrhea, 156 from children without recent diarrhea, and five lacking diarrhea data. Additionally, seven samples came from children who had received RUTF or a local alternative, independent of the intervention protocol. The cohort overall exhibited a high burden of enteric pathogens, although most pathogen-positive children were asymptomatic at the time of sample collection.

#### Kenya

Kenyan samples were obtained as extracted DNA from stool samples collected in Kisumu, Kenya as part of the TOTO study^46^. The TOTO cohort comprised children generally representative of the broader Kenyan pediatric population. Children were enrolled in the TOTO study when presenting for clinical care with symptoms of respiratory illness raising clinical suspicion for tuberculosis infection. Full study enrollment criteria and clinical characteristics of the original TOTO study have been previously published^46^. Of the 228 Kenyan samples included in our analysis, 70 (30.8%) reported receiving nutritional supplmentation with RUTF or a local alternative as part of clinical care, independent of microbiome sampling protocol.

#### Nicaragua

Nicaraguan samples were obtained as stool slurries (stored 1:1 in phosphate-buffered saline, PBS) from the *Sapovirus*-Associated Gastro-Enteritis (SAGE) study^47^, which enrolled children at risk of diarrheal disease in León, Nicarauga. Full enrollment criteria and clinical characteristics of the original SAGE study have been previously published^47^. In addition to the previously described control (n=33) and diarrheal (n=32) samples, we obtained n=391 additional stool samples collected for monthly surveillance from study participants. Non-diarrheal surveillance samples were specifically selected from children who had not experienced a diarrhea episode within the preceeding month. Only surveillance samples from children aged 2-12 months were included in our 16S microbiome analysis.

#### Pakistan

The Pakistan samples were obtained as extracted DNA from fecal samples collected as part of the Study of Environmental Enteropathy (SEEM), a prospective longitudinal cohort study, conducted in Matiari, a rural area in Sindh, Pakistan, between March 2016 and March 2019.

The SEEM study collected samples from 50 healthy control children and longitudinal samples from malnourished children (WHZ< -2), who were part of a malnourishment intervention arm.

For our analysis, we included stool samples (n=29) exclusively from the healthy control group (WHZ > 0), none of whom received nutritional interventions. Approximately 1g of stool was collected from each participant by community health workers within 10 minutes of defecation. Samples were immediately transported from field sites in 4°C cooler boxes, stored at -80°C upon arrival at the laboratory, and shipped on dry ice for subsequent analysis.

#### United States

US samples consisted of frozen solid stool or stool slurry obtained from participants enrolled in the Project HOPE 1000 study^48^ (Durham, NC, USA) and Gastrointestinal Microbiome & Allergic Proctocolitis Study (GMAP) study (Boston, MA, USA)^49^. Project HOPE 1000 aims to develop a biorepository of longitudinal samples collected from healthy infants over the first 1000 days of life, with the goal of identifying early life determinants of health. The GMAP study prospectively collected stool samples from children, some of whom subsequently developed allergies and/or proctocolitis; however, GMAP samples analyzed in this study were exclusively from infants who did not develop allergic disease. Breastfeeding status was documented at the time of each sample collection for Project HOPE 1000. For GMAP, breastfeeding status was recorded at birth and 4 months of age, but not at the exact time of stool sample collection. Therefore, infants exclusively receiving formula at 4 months of age in GMAP were presumed to not be breastfed at the time of stool collection.

### DNA Extraction

*Cambodia:* Stool samples were preserved 1:1 in Zymo DNA/RNA Shield buffer (Zymo Research, Irvine, CA) and then stored at -20°C. They were then extracted using the QIAamp 96 Virus QIAcube HT (Qiagen, Germany) protocol. 200mg of solid stool or 200uL of liquid stool were combined with 1mL of stool lysis buffer (ASL, Qiagen, Germany) and vortexed in an SK38 soil grinding tube (Bertin Corp., Rockville, MD) for five minutes (Vortex Genie 2, Scientific Industries, Bohemia, NY). Following a ten minute incubation at room temperature, the lysate was centrifuged at 12,000 g for two minutes (Thermo Fisher Scientific, Waltham, MA). 200uL of this lysate was then used for DNA/RNA extraction with the QIAamp 96 Virus QIAcube HT protocol.

*Kenya:* 1g of stool was diluted in 10mL of sterile water and emulsified with sterile glass beads. The stool slurry was then filtered. DNA was extracted using the MagAttract PowerSoil DNA EP kit (Qiagen, Germany) according to the manufacturer’s instructions.

*Pakistan*: 50mg of frozen fecal sample was extracted using phenol-chloroform extraction. Briefly, samples were ground in liquid nitrogen using a mortar and pestle. 50mg of this was then transferred to a 2mL vial. 500uL of 0.1mm diameter zirconia/silica beads were added to each sample, along with 500 µL of phenol: chloroform: isoamyl alcohol (25:24:1), 210 µL of 20% SDS and 500 µL of buffer (200 mM NaCl, 200 mM Trizma base, 20 mM EDTA). Samples underwent bead beating (Mini-Beadbeater-8; Biospec) to extract DNA. Extracted DNA was purified, (Qiaquick columns, Qiagen), then eluted in 70 µL of Tris-EDTA (TE) buffer, and then quantified (Quant-iT dsDNA broad range kit; Invitrogen). Each DNA sample was adjusted to a concentration of 1 ng/uL.

### Nicaragua and USA

The Nicaraguan and USA (Durham, Boston) stool samples were diluted 1:10 in filtered phosphate buffered saline (0.2uM filter) and then stored at -80°C. Samples were thawed on ice in an anaerobic chamber. Samples were vortexed 30s and then 250uL aliquots were taken for DNA extraction. Slurry aliquots were then randomized and extracted using the Power Soil Pro MagAttract kit (Qiagen, Germany) according to manufacturer’s instructions.

### DNA Sequencing

#### 16S rRNA Sequencing

All samples were PCR-amplified using primers targeted to the V4 region of the 16S rRNA gene using primers 515F and 806R as previously described.^50^ Kenyan samples were quantified using a Qubit dsDNA HS assay kit (ThermoFisher), pooled in equimolar concentration, and sequenced on a MiSeq sequencer (Illumina; 2x250 base pair). Cambodian, Nicaraguan, Pakistan, and USA samples were processed as previously described^50–52^, diluted to a 10nM concentration, and sequenced using an Illumina MiniSeq and a MiniSeq High Output kit (FC420-1004) with paired-end 150-bp reads.

#### FoodSeq

FoodSeq was carried out as described in previously published protocols.^53^ For plant FoodSeq, samples were amplified in a 10uL volume with 3 μL of extracted DNA template and either a) 0.5 μL each of 10 μM forward and reverse primers (IDT), 2 μL of 5X KAPA HiFi buffer, 0.3 μL of 10 mM dNTPs, 0.1 μL of 100X SYBR Green I (Life Technologies), 0.1 μL KAPA HiFi polymerase, 3.5 μL nuclease-free water, or b) 5uL of KAPA HiFi 2X, 0.3uL each of 10 μM forward and reverse primers (IDT), 0.1uL of 100X SYBR Green I (Life Technologies) and 1.3uL of nuclease-free water. Method (b) uses the same components as method (a), but in a premixed format with a slightly higher concentration of polymerase. Cycling conditions were 95°C for 3 minutes, followed by 35 cycles of 98°C for 20 seconds, 63°C for 15 seconds, and 72°C for 15 seconds.

For animal FoodSeq, samples were amplified in a 10uL volume with 1uL of extracted DNA template and 0.5uL of 10uM forward and reverse primer and 1uL of 100uM human blocking primer (IDT), 0.1 uL of 100X SYBR Green, 0.25uL of 20mg/mL bovine serum albumin, and 5uL of 2X Accustart. The amplification primers were 12SV5F and 12SV5R^54^ with Illumina overhang adapter sequences added at the 5′ end, and the human blocking primer was DeBarba14 HomoB^55^. Cycling conditions were an initial denaturation at 94°C for 3 minutes, followed by 35 cycles of 94°C for 20 seconds, 57°C for 15 seconds, and 72°C for 1 minute.

For both plant and animal FoodSeq, primary amplification was followed by secondary PCR amplification to add Illumina adapters and dual 8-bp indices for sample multiplexing. Samples were amplified in a 50uL volume using 5uL primary PCR product diluted 1:100 in nuclease free water, 25uL KAPA HiFi 2X Buffer, 0.5uL 100X SYBR Green I, 9.5uL nuclease free water, 5uL each of 2.5uM forward and reverse indexing primers.

Amplicons were cleaned (Ampure XP, Beckman Coulter, Brea, CA), quantified (QuantIT dsDNA assay kit, Invitrogen, Waltham, MA), and pooled in equimolar ratios to create a sequencing pool. If samples were too low in concentration to be pooled at a equimolar ratio, they were added up to a set volume of 15-20uL. The pooled library was then concentrated, gel purified, quantified by both fluorimeter and either qPCR or BioAnalyzer (Agilent, Santa Clara, CA). If trnL and 12SV5 amplicons were to be sequenced on the same run, they were combined at this point in equimolar amounts. Final sequencing library was spiked with 30% PhiX (Illumina, San Diego, CA) to mitigate low nucleotide diversity. Libraries were sequenced on an Illumina MiniSeq according to manufacturer’s instructions using a 150-cycle Mid or High kit, (Illumina, San Diego, CA, USA), depending on the number of samples in each pool.

### Data analysis

All packages used are available as a Singularity container hosted by the Duke Gitlabs (https://gitlab.oit.duke.edu/lad-lab).

### 16S rRNA

All code used in bioinformatic pipeline to process 16S rRNA amplicon data can be found on Github at tjk30/16S-pipeline. To summarize, following conversion to fastq format using Illumina’s bcl2fastq package, DNA sequencing results were demultiplexed using qiime2 with the --p-no-golay-error-correction flag. Trimmomatic was used to remove primer sequences. Dada2 (v1.22.0) was then used to filter, trim, merge, and assign species to reads using the SILVA database v138.1. Samples with <5000 total reads were excluded from analysis.

Richness was calculated as the sum of unique ASVs that had reads>0. Shannon diversity was calculated using estimate_richness() from the *phyloseq* package (v.1.48.0) in R (v.4.1).

Unweighted Unifrac was performed using the SILVA v138.1 SSU phylogenetic tree and the Unifrac() function from the *phyloseq* package. Ordination of sample distances was then performed using cmdscale() from the R package *stats* (v4.4.1) with the flag eig=TRUE. Envfit was run using the envfit() function from the R package *vegan (*v2.6.6.1). For calculation of the variation captured by age and country, both variables were included in the model. For the breastfeeding and birth mode models, each model was run controlling for country and age.

For microbiome clustering analysis, we excluded samples from children known to be experiencing diarrhea at the time of stool sample collection (n=33/934) or acute malnutrition (WHZ <-2), as these conditions are known to significantly impact gut microbiome composition^56,1^. Diarrhea data were available only from the Cambodian and Nicaraguan cohorts. Age was binned as follows: 0-2mo, 3-5mo, 6-8mo, 8-10mo, 11-13mo, 14-16mo,17-19mo, 20-22mo, 23-25mo, and 26+mo (binned together due to limited sampling >24mo in Cambodia and USA). Clustering was limited to 0-36 month old children to ensure that each age bin consisted of children from multiple countries, as only Kenya provided samples from children older than 36 months. Taxa were filtered to those with at least 0.5% relative abundance in at least one age bin. Rare taxa, defined as taxa detected in two or fewer samples, were removed from analyses.

The samples that passed filtering criteria (n=818) were then used to calculate the average relative abundance of each ASV within each age bin. In order to normalize across taxa with different relative abundance ranges across age, relative abundances for each taxon were converted to z-scores. Clustering analysis was performed on the Euclidean distances of the taxa z-scores using Unweighted Pair Group Method with Arithmetic Mean (UPGMA; implemented with the hclust() function from the R package *stats*, method=’average’). Clustering analysis was then used to generate an k=10 clusters using the *stats* function cutree().

After identifying microbiome clusters in the subset of healthy children, we reincorporated the excluded "sick" samples back into the dataset for subsequent analyses, as we wanted to comprehensively evaluate the identified microbial clusters within the context of broader childhood conditions and maximize statistical power. We calculated the cluster abundances for Clusters 1-10 for all samples in the full dataset (n=934) by merging the taxa identified within each cluster and then performing centered log ratio (clr) normalization using the R package microbiome (v1.26.0).

### FoodSeq

All code and databases used in the bioinformatic pipeline to process FoodSeq amplicon data can be found on Github at LAD-lab/mb-pipeline. Bioinformatic processing was conducted as previously described^30^. To summarize, following conversion to fastq format and demultiplexing using Illumina’s bcl2fastq package, primer sequences were trimmed using the qiime2 cutadapt wrapper with the following flags: --p-error-rate 0.15, --p-minimum-length 1, --p-overlap 5, p-discard-untrimmed. Paired reads were then merged using the qiime2 dada2 wrapper with the following flags: --p-trunc-len-f 0, --p-trunc-len-r 0, --p-max-ee-f 2, --p-max-ee-r 2, --p-trunc-q 2, p-min-overlap 12, --p-pooling-method ’independent’.

As we used a non-error-correcting polymerase for the 12SV5 PCR amplification reactions to maximize human blocking primer efficiency (resulting in more error-prone amplification), ASVs mapping to the same species were glommed using tax_glom() in the R package *phyloseq* prior to analysis. This meant that ASVs were assigned to the most similar species in the database used from Schneider et al.^57^

pFR was calculated as the sum of unique assigned ASVs that had any detectable reads. Unassigned ASVs that did not match a human food in our database were excluded from pFR. Samples with a pFR >5 standard deviations outside the median were excluded from further analysis.

For food clustering analysis, foods that did not have at least one read in one sample were removed and a matrix was generated of the percent of samples in each country where each assigned ASV was detected. Clustering was performed on the Euclidean distance between each food based on its prevalence in the 5 countries using the “complete” method in the function hclust() from the R package *stats*. The R package *ggtree* was then used for dendrogram visualization.

### Statistical Analysis

#### Data Ordinations

To account for the compositional nature of the data, we calculated the centered-log ratio (CLR) transformation of DNA sequencing read counts using the transform() function of *microbiome* v1.26.0. Principal component analysis (PCA) was then performed on CLR-transformed read counts using the prcomp() function in the *stats* R package (v4.4.1).

#### Linear model design

Generalized linear models were used to model the relationship between dietary factors and microbiome outcomes. We included age as a covariate in our models due to the strong relationships between age and our microbiome outcomes of interest established in previous literature.^1^ We further controlled for country due to differences in age distribution, malnutrition, enteric pathogen exposure, and breastfeeding duration between the countries assessed. Our choice to include interaction terms in our models was determined by choosing the model with the lowest AIC and significant chi-squared p-value in a likelihood ratio test between the reduced model and saturated model.

The relationship between dietary diversity and microbial diversity was modeled using the call *glm(microbiome_diversity ∼ pFR + country + age_months + country:age_months + pFR:country, family = ’gaussian’)*. The relationship between dietary PC1 and microbiome clusters was modeled using the call *glm(cluster ∼ age_months + country + country:age_months + PC1 + country:PC1 + age_months:PC1, family = ’gaussian’)*. Cluster associations with subsequent PCs were not tested, as country was highly collinear with PC2, accounting for half of its variance (ANOVA, p < 2e–16; R² ≈ 0.50 for country), which would render PC2 effects indistinguishable from country-level associations.

When fitting within-country models, the following models were used, depending on available covariate data:

USA and Kenya (breastfeeding data not available):

*glm(microbiome_outcome ∼ diet_variable + age_months, family = ’gaussian’)*

Pakistan: (lacked breastfeeding data; all samples came from one timepoint (12-15 months):

*glm(microbiome_outcome ∼ diet_variable, family = ’gaussian’)*

Cambodia and Nicaragua (breastfeeding data available):

*glm(microbiome_outcome ∼ diet_variable + currently_breastfed + age_months + currently_breastfed:age_months + diet_variable:currently_breastfed, family= ’gaussian’)*

When accounting for the effects of WHZ on microbiome richness, the following models were used:

Kenya:

glm(microbiome_diversity ∼ WHZ + pFR + age_months + sex)

Cambodia:

*glm(microbiome_diversity ∼ WHZ + pFR + age_months + sex + currently_breastfed +*

*pFR:currently_breastfed + currently_breastfed:age_months + WHZ:currently_breastfed)*

### Institutional and Ethical Review

Stool and extracted stool DNA samples were obtained through collaborations with existing studies via an IRB exemption (Pro00100567) from the Instutitional Review Board at Duke University.

Use of Kenyan pediatric stool samples in this analysis was reviewed and approved by the institutional review boards of the US Centers for Disease Control and Prevention, the Kenya Medical Research Institute, and the Jaramogi Oginga Odinga Hospital. Written informed consent for the use of these stool samples was given by parents or legal guardians of participants.

### U.S. CDC Disclaimer

This work has been supported in part by the President’s Emergency Plan for AIDS Relief through the U.S. Centers for Disease Control and Prevention, The findings and conclusions in this report are those of the authors and do not necessarily represent the official position of the funding agencies.

## Data Availability

All data produced in the present study are available upon reasonable request to the authors

## Acknowledgements

Nicaraguan samples: The SAGE study was made possible with the NIH grants R01AI127845 and: K24AI141744.

Kenyan samples: We acknowledge the Duke Microbiome Core Facility for performing the DNA extractions, quality control, and preparing the 16S rRNA gene sequencing library; the Duke Sequencing and Genomic Technologies Shared Resource for sequencing the 16S rRNA gene library. We also thank the Duke Microbiome Core Facility for performing DNA BioAnalyzer quality assessments.

Pakistan samples: SEEM study was a multi-national collaboration between Aga Khan University (Pakistan), Washington University in St. Louis, the University of Virginia, and Cincinnati Children’s Hospital Medical Center, funded by the Bill and Melinda Gates Foundation (2016-2019). Marty Meier provided technical assistance in stool DNA purification.

Linda Adair, Jun Zeng, Brianna Petrone, Michelle Kirtley, provided valuable scientific guidance to this study.

Finally, we thank all of the children who were participants in this study, as well as their parents. Without their time, effort, and participation, this study would not have occurred.

---

**Fig. S1.**
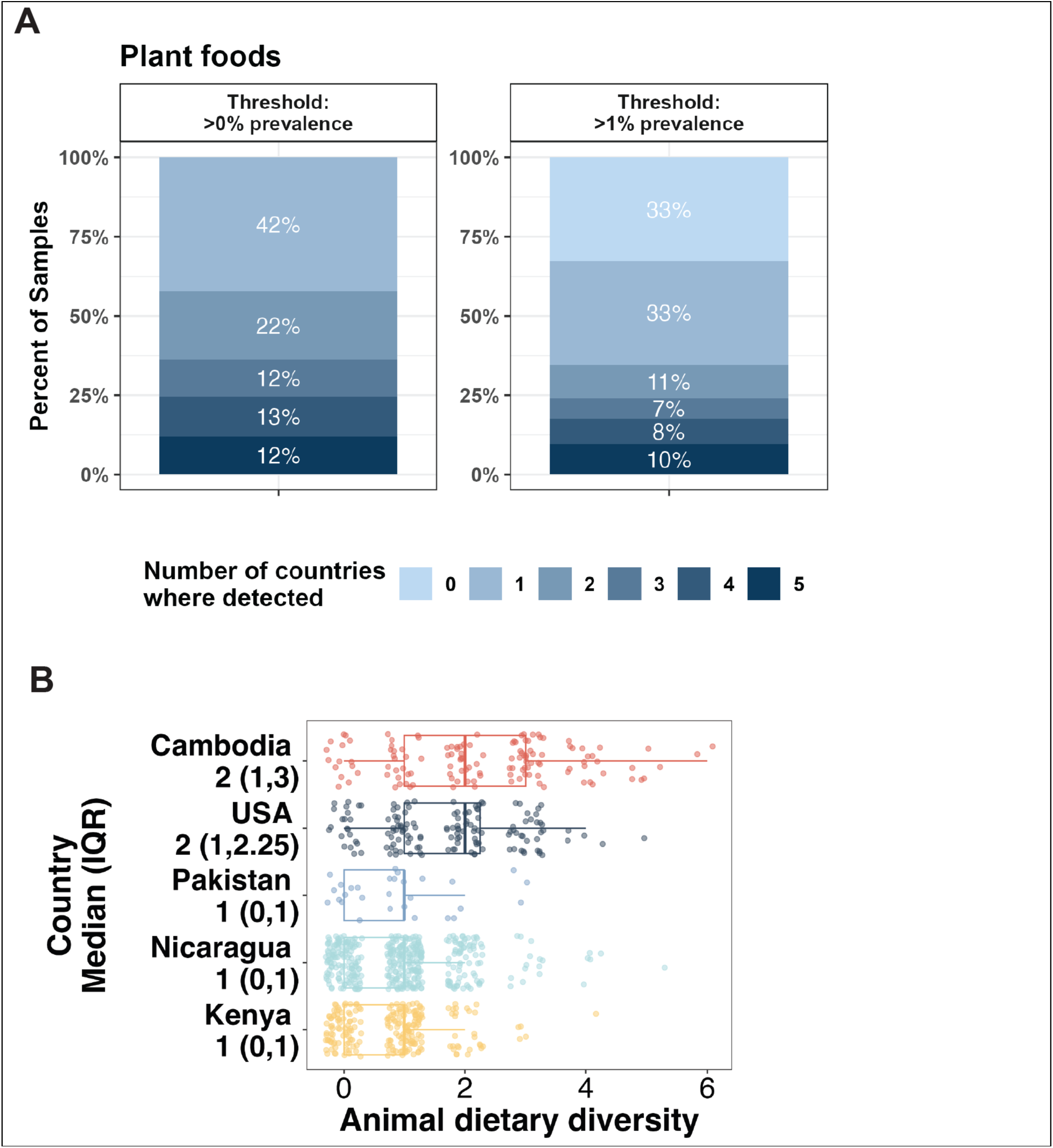
(A) Distribution of foods by how many countries they were each detected in. A food was counted as ‘detected’ if it was detected in at least 1 sample in that country (>0% threshold), or at least 1% of samples in that country (>1% threshold). (B) Animal FoodSeq diversity by country.

**Fig. S2.**
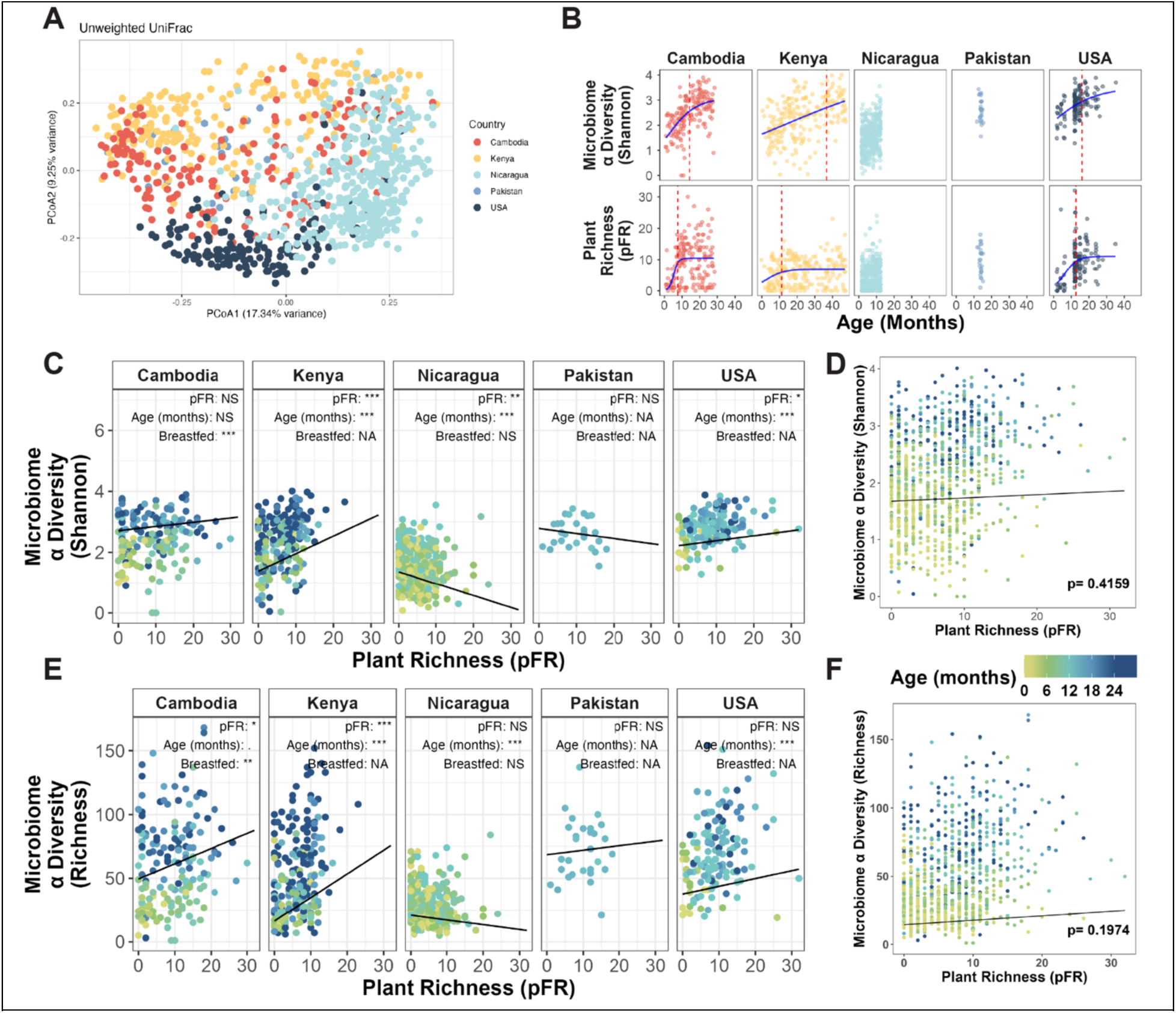
**(A)** Unweighted Unifrac ordination. **(B)** Plant and microbiome diversity versus age, with Pakistan and Nicaragua included (see: Fig. 3B). **(C)** Scatterplots showing relationships between microbial Shannon diversity and pFR within country. (D) Scatterplot showing relationship between microbial Shannon diversity and pFR. (E) Scatterplot showing relationships between microbial richness and pFR. For C-E, line plots the coefficient of pFR derived from a generalized linear model controlling for age (C-E), and, where applicable, country (D-E) and breastfeeding status (C, Cambodia and Nicaragua).

**Fig. S3:**
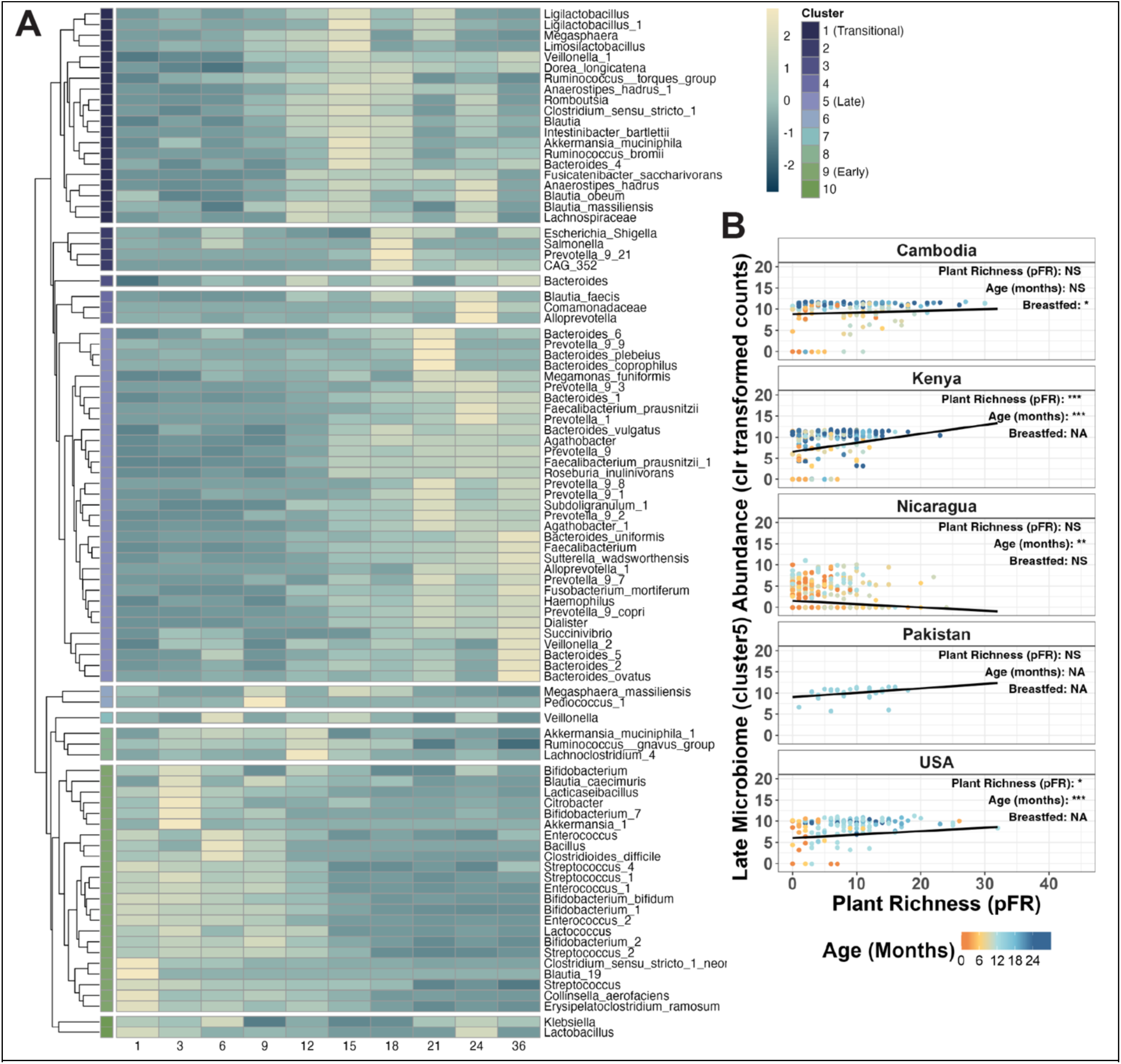
**(a)** Heatmap plotting age-differential microbiome clusters. **(b)** Late microbiome abundance correlated with pFR within country while controlling for breastfeeding status, where available, and age. Similar to microbiome diversity, late microbiome abundance is positively correlated with plant richness in Kenya and the USA.

**Fig. S4.**
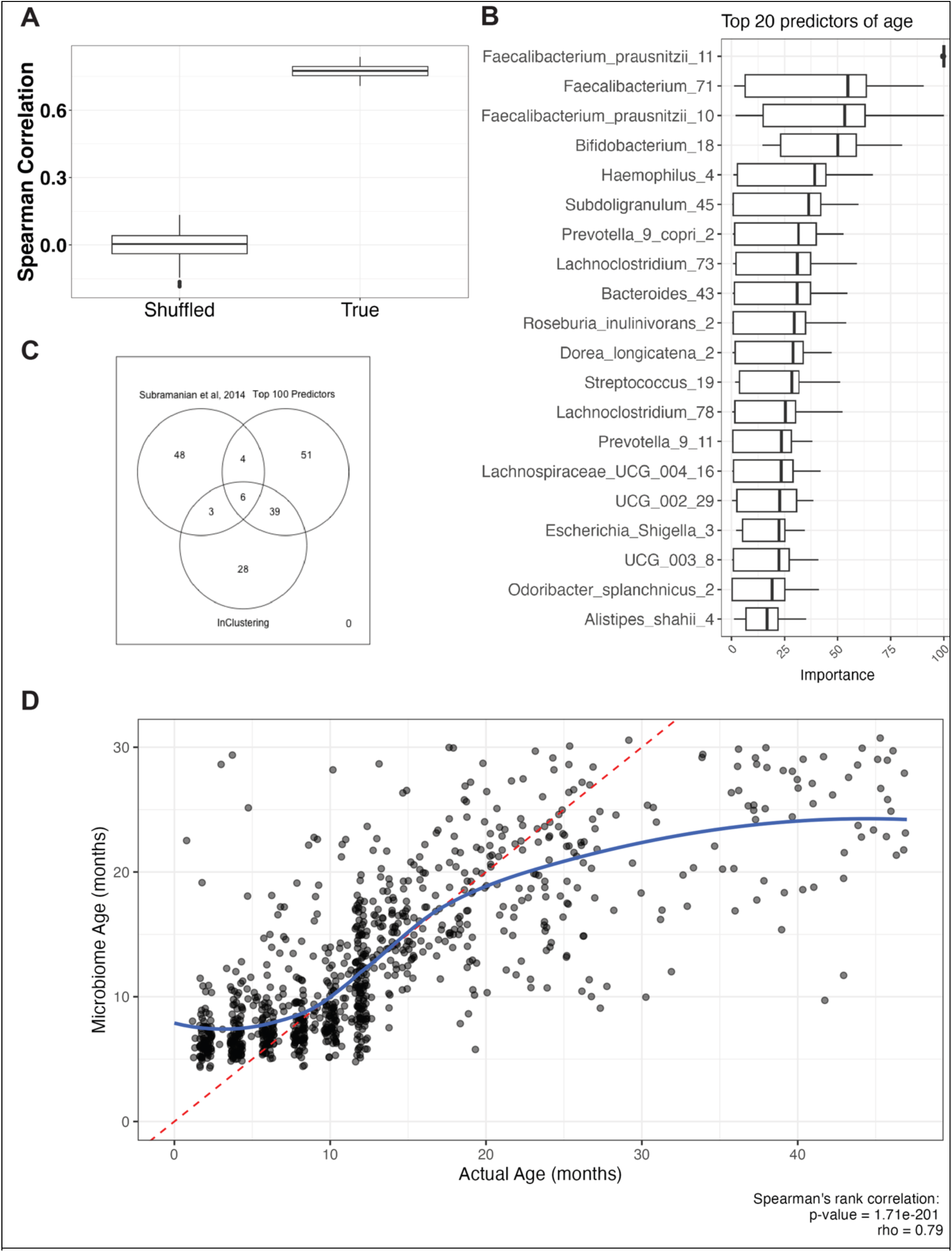
(A) Model fit for classifier predicting age from shuffled or true 16S rRNA relative abundance data. (B) Top predictors from Random Forest model. (C) Exact sequence matches between the top 100 predictors in the Random Forest model, the taxa identified in the Early, Transitional, and Late clusters, and a previous study that first employed this analysis. (D) Plot of microbiome age versus actual age. Red dashed line is a theoretical perfect concordance between the two.

**Fig. S5.**
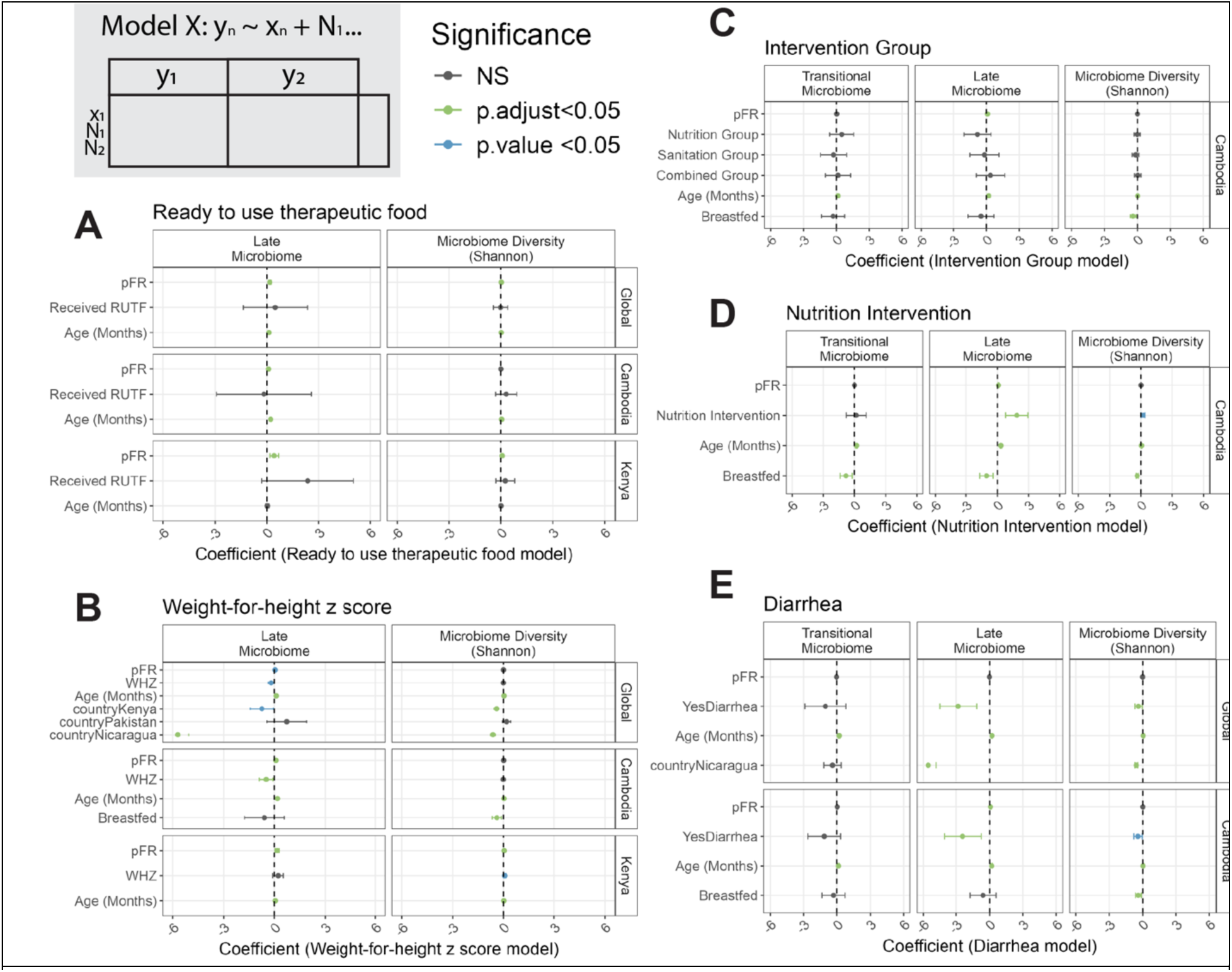
Generalized linear models controlling for country-specific variables. **(A)** Models controlling for RUTF consumption. **(B)** Models controlling for weight-for-height z score (WHZ). **(C)** Models controlling for study intervention group within the Cambodian cohort. **(D)** Models controlling for whether or not Cambodian children were in an intervention group that included a nutrition intervention. **(E)** Models controlling for diarrhea. Each column represents an outcome variable, and each row represents a predictor variable. Results are grouped by whether the test was run on all samples with non-NA values for that variable (‘Global’) or within a specific country. Missing countries did not collect data for that variable.

**Fig. S6.**
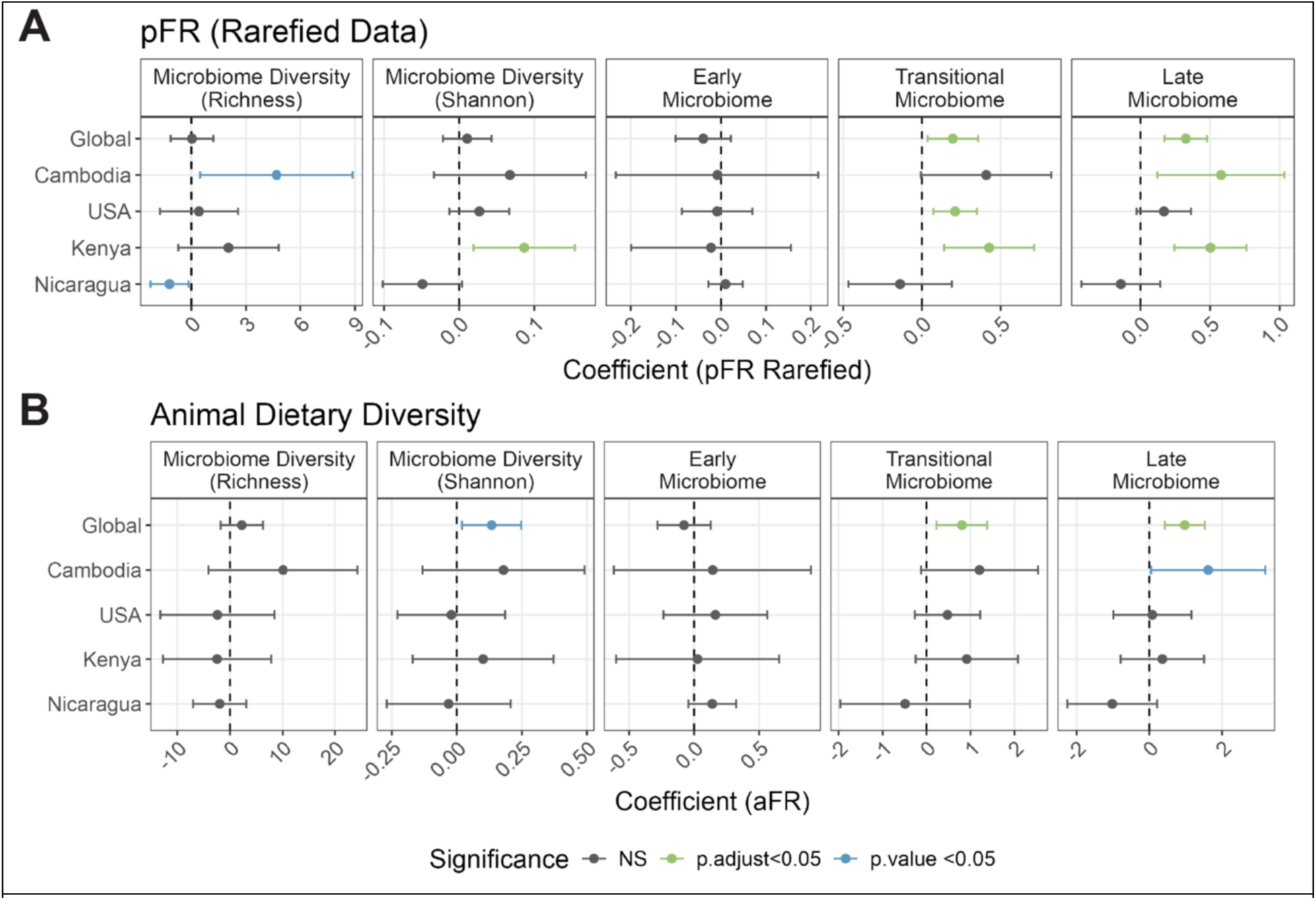
Summary of generalized linear model results for other diet metrics. **(A)** Model results for pFR calculated from data rarefied to 500 reads. **(B)** Model results for animal dietary diversity (aFR). Y axis is the dataset the test was run on (‘global’ for all samples, or within a specific country.) Pakistan was omitted due to low sample size (n=29). X axis represents the coefficient, indicative of effect size, of that variable in the model.

**Table S1:** Cohort Characteristics.

| Table S1: Cohort Characteristics | Cambodia | Kenya | Nicaragua | Pakistan | USA |
| --- | --- | --- | --- | --- | --- |
| Samples | 178 | 228 | 457 | 29 | 144 |
| Participants | 178 | 228 | 141 | 29 | 132 |
| Age range of cohort (months) | 1-28 | 0-47 | 2-25 | 12-15 | 2-35 |
| Age, Months (mean, SD) | 14.31 ,7.47 | 22.44, 13.28 | 7.33, 4.09 | 13.86, 0.58 | 13.42, 5.84 |
| Male (%) | 95 (53.4) | 111 (48.7) | 227 (49.7) | 17 (58.6) | 76 (52.8) |
| WHZ (mean (SD)) | -0.69 (0.98) | -0.92 (1.38) | 0.64 (1.19) | -0.15(0.66) | NaN (NA) |
| HAZ (mean (SD)) | -1.09 (2.35) | -0.42(2.41) | 0.18 (1.20) | -0.99(1.21) | NaN (NA) |
| WAZ (mean (SD)) | -1.11 (1.47) | -0.86 (1.82) | 0.55 (1.02) | -0.59(0.91) | NaN (NA) |
| Received nutrition (RUTF or local alternative) | 7 (4%) | 70 (30.8%) | NA | NA | NA |
| National Human Development Index<sup>58</sup> | Low | Medium | Medium | Low | Very High |
| Urbanization | Rural | Urban | Urban | Rural | Urban/Suburban |
| Currently Breastfed (%) |  |  |  |  |  |
| Don't Know | 9 (5.1) | 228 (100.0) | 0 (0.0) | 29 (100.0) | 71 (49.3) |
| No | 69 (38.8) | 0 (0.0) | 133 (29.1) | 0 (0.0) | 54 (37.5) |
| Yes | 100 (56.2) | 0 (0.0) | 324 (70.9) | 0 (0.0) | 19 (13.2) |
| pFR (mean (SD)) | 9.37 (6.69) | 6.20 (4.30) | 4.48 (3.88) | 9.90 (4.44) | 8.90 (5.71) |
| Diarrhea (%) |  |  |  |  |  |
| Don't Know | 5 (2.8) | 228 (100.0) | 392 (85.8) | 29 (100.0) | 144 (100.0) |
| No | 159 (89.3) | 0 (0.0) | 33 (7.2) | 0 (0.0) | 0 (0.0) |
| Yes | 14 (7.9) | 0 (0.0) | 32 (7.0) | 0 (0.0) | 0 (0.0) |

